# Association between Covid-19 BNT162b2 vaccination or SARS-CoV-2 infection and immune mediated diseases in 5 million 5-to-19-year-olds in four Nordic countries

**DOI:** 10.1101/2025.03.09.25323620

**Authors:** German Tapia, Rickard Ljung, Emilia Myrup Thiesson, Nicklas Pihlström, Tuomo Nieminen, Inger Johanne Landsjøåsen Bakken, Hanne Løvdal Gulseth, Hanna Nohynek, Øystein Karlstad, Anders Hviid

## Abstract

**Background:** We investigated new onset of autoimmune hepatitis (AH), Guillain-Barré syndrome (GBS) and Type 1 Diabetes (T1D), and potential increased disease activity (measured as recurrent hospital visits) of Juvenile Rheumatoid Arthritis (JRA), Multiple Sclerosis (MS) and T1D, following BNT162b2 vaccination or SARS-CoV-2 infection in children and adolescents.

**Methods:** We used nationwide data from Norway, Denmark, Finland, and Sweden, using 28- and 180-days risk periods following BNT162b2 vaccination or a positive test for SARS-Cov-2 infection. Individuals aged 5-19 years during the study period (1^st^ January 2021-31^st^ December 2022) were included. We used a common data model, using individual level data linked from nation-wide registers and a shared analysis script to harmonize data and analysis. Poisson regression was used to estimate relative risks (RRs) comparing outcome rates in risk windows to unvaccinated or uninfected follow-up rates. This was supplemented with self-controlled case series analyses. Results from different countries were analyzed jointly in a meta-analysis.

**Results:** We observed no clear association between BNT162b2 vaccination and new-onset AH, GBS or T1D. Neither did we observe any clear association with recurrent hospital visits following BNT162b2 vaccination or SARS-CoV-2 infection. An association between SARS-CoV-2 infection and GBS was observed in both the 28-day and 180-day risk window (relative risk (RR) 15.10, 95% CI, 1.11-205.94, and RR 3.85, 1.33-11.14, respectively).

**Conclusions:** BNT162b2 vaccination was not associated with new-onset or recurrent hospital visits of these diseases in children and adolescents. Sars-CoV-2 infection was associated with new-onset GBS. Estimates were however uncertain due to the small number of cases.

## INTRODUCTION

Several immune-mediated diseases have been associated with childhood infections and vaccinations.^1–3^ Although the mechanisms are unknown, the involvement of the immune system in these diseases makes potential associations plausible. SARS-CoV-2, or vaccines against SARS-CoV-2, could therefore potentially be associated with development of immune-mediated diseases, or increased disease activity of these.

Present studies during the COVID-19 pandemic have reported that infection in children and adolescents is mostly mild.^4,5^ Clinical trials of vaccinations in children and adolescents have shown good efficacy and short-term safety.^6–9^ There are presently few studies on adverse events following COVID-19 vaccination or infection in children and adolescents,^10–12^ and most lack sufficient follow-up to detect non-acute associations. There have been reports of Autoimmune Hepatitis (AIH),^13–15^ Guillain-Barré Syndrome (GBS),^16–18^ Type 1 Diabetes (T1D),^14,19^ Juvenile Rheumatoid Arthritis (JRA),^20^ and Multiple Sclerosis (MS) following vaccination or infection in adults.^21^ As many immune-mediated diseases are believed to have a long pre-clinical period, a long follow-up is needed to evaluate the safety of vaccines with respect to immune-mediated diseases.

There is a need for studies investigating long-term or rare adverse events after vaccination and infection, both to build public trust in the vaccination programs, and for risk-benefit analyses of vaccination in children and adolescents. In this study, we aimed to investigate potential associations between SARS-CoV-2 vaccination or infection and selected immune-mediated diseases (both new onset, and increased disease activity [measured as recurrent hospital visits]) in children/adolescents aged 5 to 19 years in the Nordic countries.

## MATERIAL AND METHODS

This study publishes results of an investigation initiated by the European Medicines Agency (EMA). The overall aim was to study possible associations between COVID-19 vaccination and infection and selected immune-mediated diseases in four Nordic countries (Denmark, Finland, Norway, Sweden), as these might be too rare in children/adolescents to study in a single country. The participating countries implemented national vaccination campaigns against SARS-CoV-2 and provided free vaccinations to all residents but had some differences in distribution plans and recommendations for children and adolescents.

### Ethics

Due to privacy concerns, we do not present results for subpopulations with <5 cases. Data was obtained and analyzed following the country-specific laws and ethical regulations, all conforming to the principles embodied in the Declaration of Helsinki. For country-specific details see the online supplement.

### Study design

We included all residents 5-19 years of age in each country (Sweden: 12-19 years, as Sweden did not vaccinate children <12 years of age during the study). The study period started 1 January 2021 and ended 31 December 2022. Using unique personal identifiers, each country linked individual-level data on demographic variables, vaccination dates, infection dates, outcomes, and relevant covariates from nationwide registers.^22^

### Outcomes

Five immune-mediated diseases were studied: Autoimmune Hepatitis (AIH), Guillain-Barré Syndrome (GBS), Type 1 Diabetes (T1D), Juvenile Rheumatoid Arthritis (JRA), and Multiple Sclerosis (MS) (Supplemental Table 1). These were selected on the basis of a suspected link to SARS-CoV-2 vaccination or to childhood vaccinations. We investigated both incident disease (for AIH, GBS and T1D) and recurrent hospital visits (for MD, JRA and T1D). Recurrent hospital visits were investigated to determine if disease flares, or increased disease activity, followed SARS-CoV-2 infection or vaccination. New onset/incident cases were defined as the first occurrence in the study period of a diagnosis without preceding occurrences in the 2-year period 1 January 2019–31 December 2020. When analyzing recurrent hospital visits, we used a wash-out period of 30 days following a hospital visit before counting a new disease occurrence. It should be noted that for many immune-mediated diseases, recurrent hospital visits as defined above might not necessarily represent sudden disease activity but could be onset of complications, or routine check-ups.

### Exposures

BNT162b2 vaccination was the primary exposure, due to its almost exclusive use in adolescents and children in this setting. We present data on one, two and three doses of BNT162b2. We do not present data on other vaccines (such as mRNA-1273, or vector vaccines AZD1222 and AD26.COV2.S) due to their marginal use and the rarity of outcomes in the study population. We analyzed up to the third dose, presenting results per dose. The third dose was mainly administered in 16-19-year-olds and individuals at high risk for serious SARS-CoV-2 infections. Individuals were censored upon receiving a fourth dose of BNT162b2, a mRNA-1273 vaccine, or an adenoviral vector vaccine.

SARS-COV-2 infection, defined by the first positive RT-PCR test, was a secondary exposure. We treated SARS-COV-2 infection as a censoring event for the evaluations of BNT162b2 vaccination as an exposure. SARS-COV-2 vaccination was a censoring event when analyzing infection as an exposure, as were reinfections. Vaccination and infection were included as time-varying exposures.

We included both a short-term (0-27 days after exposure), and a long-term risk period (0-179 days after exposure). Most other studies have operated with a risk period of around 4-6 weeks, which is sufficient for an acute onset. We chose 0-179 days based on expected follow-up in our study while including a reasonable lag for pathogenesis or diagnosis.

### Covariates

We included the following potential confounders: age, sex, calendar year and month, region of residence (country-specific), mother’s birth country (Nordic, Western, non-Western; not available in Finland), comorbidities (asthma, other chronic respiratory diseases, chronic cardiac disease, renal disease, epilepsy or convulsions, congenital malformations and chromosomal abnormalities, malignancy or immunodeficiency and psychiatric disorder), and high-risk group (country-specific, used to identify children at high risk of serious SARS-CoV-2 infection outcomes). ICD-10 codes of the comorbidities are shown in Supplemental Table 1.

### Statistical methods

We used two different designs in the present study, analyzing data in both a cohort design and a self-controlled case series (SCCS) analysis. We considered a P-value <0.05 as statistical significance, or evidence of an association. If both the cohort and SCCS estimates had P-values <0.05 (and estimates were not contradictory) we considered the association consistent. We focused on reporting the estimates for those exposure-outcome pairs for which evidence of association was found.

For the cohort analysis, adjusted incidence rate ratios were estimated by comparing the risk period follow-up time with unexposed follow-up time. We used multivariate Poisson regression on the outcome counts with the logarithm of the follow-up time as the offset. For the self-controlled case series analysis,^23,24^ we compared the risk period follow-up to the unvaccinated period to estimate incidence rate ratios using conditional Poisson regression with direct adjustment for calendar year and month. We used a 14-day pre-risk period before vaccination to account for a potential healthy vaccinee effect. The pre-risk period of interest was not included in the unvaccinated reference period and was included before any dose. An advantage of the self-controlled method is that all time-invariant confounders are accounted for, but it has the assumption that outcome does not influence future risk of exposure.

In the analysis of recurrent hospital visits, each individual could contribute with multiple hospital visits during follow-up. The follow-up was not censored in the cohort analyses in the event of a relevant hospital visit occurring and we did not only consider the first recording of the immune-mediated disease diagnosis in the SCCS analyses.

Our main analysis included the whole study population, with preplanned sensitivity analyses stratifying for sex and age-groups (5-11, 12-15 and 16-19 years). Sensitivity analyses were planned to describe potential associations in the main analysis further and were *a priori* not considered to be interpreted as evidence for an association if no association was observed in the main analysis. We used a common data model (CDM) and a shared analysis script written in R version 4.2.2 to harmonize data structure and analysis, available by reasonable request. Results from different countries were analyzed jointly in a random effects model meta-analysis, using the *mixmeta* R package. We did not adjust for multiple testing, as we expected chance findings to not replicate across different countries.

## RESULTS

In total, 5,029,084 5-to-19-year-olds (49% girls) were included, from Denmark (n=1,035,158), Finland (n=1,018,113), Norway (n=1,094,157) and Sweden (n=1,881,656). The total number of BNT162b2 doses administered was 4,145,463, with 2,387,657 individuals vaccinated at least once. Denmark had highest coverage (65.3%), followed by Finland (52.6%), Norway (40.1%) and Sweden (39.2%). All countries had two doses as the most common schedule, except Norway with a single dose being most common. Most 16-19-year-olds and 12-15-years-olds were vaccinated (85.1%, and 74.5%, respectively), while few 5-11-year-olds were (14.5%). Infections were reported for the highest proportion of the population in Denmark (71.0%), followed by Norway (42.5%), Sweden (23.5%) and Finland (21.4%). Vaccine uptake and SARS-CoV-2 infections did not differ by sex, but children with comorbidities had higher vaccine uptake and slightly lower infection rates. For details, see Table 1.

**Table 1:**
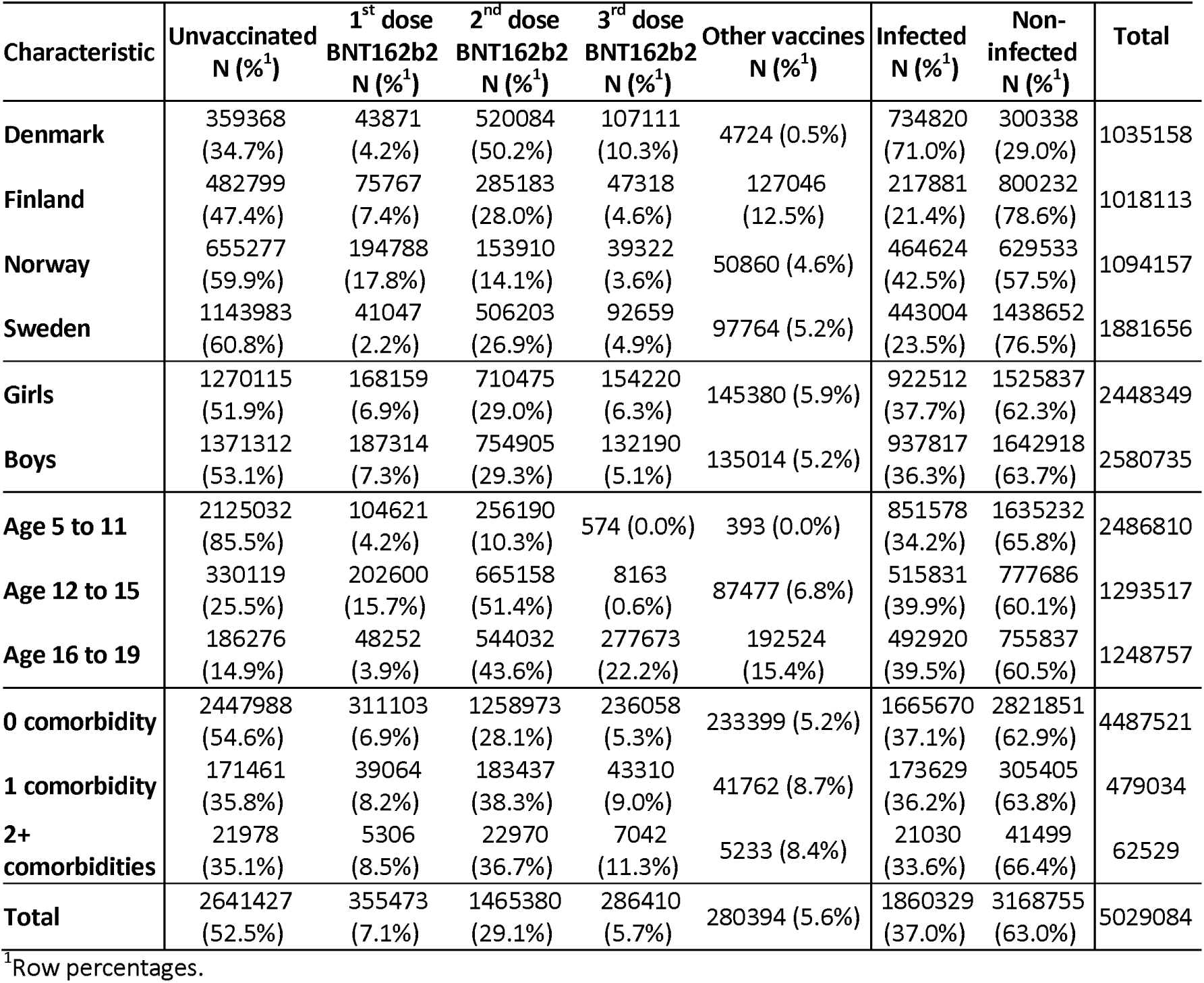
Vaccination and infection status at study end among 5-to-19-year-olds in Denmark, Finland, Norway, and Sweden.

### New-onset Results

The incidence rates for AIH, T1D and GBS, stratified by age group, are shown in Table 2. There were few exposed – vaccinated or infected – cases of AIH, and GBS, particularly in the 28-day risk period (Table 3). There was no evidence of association between vaccination and AIH, T1D or GBS in the 28- or 180-day risk period, except for 3^rd^-dose BNT162b2 vaccination and GBS in the cohort analysis of the 180-day risk period. This observation should be interpreted cautiously as it was based on very few cases, resulting in very imprecise estimates (Relative Risk [RR] 20.93, 95%CI 1.02-431.13) (Table 3). SARS-CoV-2 infection associated with GBS, both in the 28- (RR 15.10, 95%CI 1.11-205.94) and 180-day (RR 3.85, 95%CI 1.33-11.14) risk period, but the estimates were imprecise due to few cases and should be interpreted cautiously (Table 3).

**TABLE 2:** Incidence rates (per 100,000 person-years of follow-up) of onset of immune-mediated diseases.

| New-Onset Event | Age 5 to 11 |  |  | Age 12 to 15 |  |  | Age 16 to 19 |  |  | All |  |  |
| --- | --- | --- | --- | --- | --- | --- | --- | --- | --- | --- | --- | --- |
|  | Number of cases | Person-years of follow-up | Incidence rate (IR) | Number of cases | Person-years of follow-up | Incidence rate (IR) | Number of cases | Person-years of follow-up | Incidence rate (IR) | Number of cases | Person-years of follow-up | Incidence rate (IR) |
| Autoimmune hepatitis | 15 | 3479828 | 0.4 | 24 | 1845043 | 1.3 | 41 | 1689633 | 2.4 | 80 | 7014504 | 1.1 |
| Guillain-Barré syndrome | 16 | 3479767 | 0.5 | 14 | 1845114 | 0.8 | 14 | 1689761 | 0.8 | 44 | 7014642 | 0.6 |
| Type 1 diabetes | 1209 | 3474016 | 34.8 | 1035 | 1835040 | 56.4 | 657 | 1677523 | 39.2 | 2901 | 6986579 | 41.5 |
| **Flares** |  |  |  |  |  |  |  |  |  |  |  |  |
| Juvenile arthritis | 7597 | 2170.0 | 3.5 | 10174 | 3361.1 | 3.0 | 9009 | 4107.2 | 2.2 | 26780 | 9638.3 | 2.8 |
| Multiple sclerosis | 8 | 1.4 | 5.8 | 135 | 27.9 | 4.8 | 420 | 99.7 | 4.2 | 563 | 129 | 4.4 |
| Type 1 diabetes | 18869 | 3363.6 | 5.6 | 31858 | 6733.9 | 4.7 | 33541 | 9080.2 | 3.7 | 84268 | 19177.4 | 4.4 |

**Table 3:** Relative risks of new onset of immune-mediated diseases.

|  | SCCS |  | Cohort |  |  |
| --- | --- | --- | --- | --- | --- |
| 28 days Risk Period | Number of cases | Relative Risk (95% CI) | Number of cases | Relative Risk (95% CI) | Countries included<sup>1</sup> |
| Autoimmune hepatitis |  |  |  |  |  |
| Unvaccinated | 54 | 1 (ref) | 54 | 1 (ref) | - |
| 1<sup>st</sup> dose BNT162b2 | 0 | NA | 0 | NA | NA |
| 2<sup>nd</sup> dose BNT162b2 | <5 | 0.71 (0.07-7.45) | <5 | 1.06 (0.13-8.77) | SE |
| 3<sup>rd</sup> dose BNT162b2 | 0 | NA | 0 | NA | NA |
| Non-infected | 54 | 1 (ref) | 54 | 1 (ref) | - |
| SARS-CoV-2 infection | <5 | 1.02 (0.09-11.56) | <5 | 4.58 (0.52-40.40) | NO |
| Guillain-Barré syndrome |  |  |  |  |  |
| Unvaccinated | 31 | 1 (ref) | 31 | 1 (ref) | - |
| 1<sup>st</sup> dose BNT162b2 | 0 | NA | 0 | NA | NA |
| 2<sup>nd</sup> dose BNT162b2 | 0 | NA | 0 | NA | NA |
| 3<sup>rd</sup> dose BNT162b2 | 0 | NA | 0 | NA | NA |
| Non-infected | 30 | 1 (ref) | 31 | 1 (ref) | - |
| SARS-CoV-2 infection | <5 | 52.84 (5.70-489.40) | <5 | 15.10 (1.11-205.94) | DK, FI |
| Type 1 diabetes |  |  |  |  |  |
| Unvaccinated | 2112 | 1 (ref) | 2157 | 1 (ref) | - |
| 1<sup>st</sup> dose BNT162b2 | 80 | 0.81 (0.62-1.05) | 80 | 1.10 (0.87-1.39) | DK, FI, SE, NO |
| 2<sup>nd</sup> dose BNT162b2 | 51 | 0.66 (0.46-0.95) | 51 | 1.01 (0.76-1.35) | DK, FI, SE, NO |
| 3<sup>rd</sup> dose BNT162b2 | <5 | 0.49 (0.12-2.10) | <5 | 1.00 (0.24-4.06) | DK, SE |
| Non-infected | 2132 | 1 (ref) | 2158 | 1 (ref) | - |
| SARS-CoV-2 infection | 41 | 1.29 (0.91-1.82) | 41 | 1.31 (0.95-1.80) | DK, FI, SE, NO |
| 180 days Risk Period |  |  |  |  |  |
| Autoimmune hepatitis |  |  |  |  |  |
| Unvaccinated | 54 | 1 (ref) | 54 | 1 (ref) | - |
| 1<sup>st</sup> dose BNT162b2 | 12 | 0.67 (0.25-1.78) | 12 | 0.83 (0.37-1.86) | DK, SE, NO |
| 2<sup>nd</sup> dose BNT162b2 | <5 | 0.41 (0.03-5.00) | <5 | 1.04 (0.12-9.36) | SE |
| 3<sup>rd</sup> dose BNT162b2 | <5 | 0.73 (0.09-5.70) | <5 | 4.97 (0.83-29.85) | DK, FI, SE |
| Non-infected | 54 | 1 (ref) | 54 | 1 (ref) | - |
| SARS-CoV-2 infection | <5 | 0.43 (0.07-2.57) | 6 | 2.38 (0.87-6.56) | DK, FI, NO |
| Guillain-Barré syndrome |  |  |  |  |  |
| Unvaccinated | 31 | 1 (ref) | 31 | 1 (ref) | - |
| 1<sup>st</sup> dose BNT162b2 | 6 | 1.28 (0.27-6.17) | 6 | 1.31 (0.38-4.51) | DK, SE, NO |
| 2<sup>nd</sup> dose BNT162b2 | 0 | NA | 0 | NA | NA |
| 3<sup>rd</sup> dose BNT162b2 | 0 | NA | <5 | 20.93 (1.02-431.13) | NO |
| Non-infected | 30 | 1 (ref) | 31 | 1 (ref) | - |
| SARS-CoV-2 infection | 6 | 18.87 (1.60-223.04) | 8 | 3.85 (1.33-11.14) | DK, FI, SE, NO |
| Type 1 diabetes |  |  |  |  |  |
| Unvaccinated | 2112 | 1 (ref) | 2157 | 1 (ref) | - |
| 1<sup>st</sup> dose BNT162b2 | 433 | 0.74 (0.59-0.94) | 433 | 1.11 (0.97-1.28) | DK, FI, SE, NO |
| 2<sup>nd</sup> dose BNT162b2 | 70 | 0.57 (0.39-0.83) | 70 | 1.12 (0.75-1.69) | DK, FI, SE, NO |
| 3<sup>rd</sup> dose BNT162b2 | 20 | 0.48 (0.28-0.81) | 20 | 1.30 (0.82-2.07) | DK, FI, SE, NO |
| Non-infected | 2132 | 1 (ref) | 2158 | 1 (ref) | - |
| SARS-CoV-2 infection | 189 | 0.92 (0.73-1.15) | 189 | 0.94 (0.80-1.11) | DK, FI, SE, NO |

<sup>1</sup>Indicates which countries were included in the analysis.
NA: Not available, meaning that there is no estimate, or not available in any country. - : indicates the reference category, which includes all countries;

There was no evidence of associations when stratifying on age groups or sex, with two exceptions: the observation of increased risk of GBS following SARS-CoV-2 infection was driven by the 5-11-year olds, and the risk of T1D following SARS-CoV-2 infection was increased in 16-19-year-olds (Supplemental Table 2)

### Recurrent Hospital Visits

Characteristics of individuals contributing to the analysis of recurrent hospital visits are shown in Supplemental Table 3. The incidence rates, stratified by age group, for JRA, MS and T1D are presented in Table 2. Younger age groups had more visits than the older groups (Table 2). Almost all hospital visits were outpatient contacts, with the most common number of visits being three (Figure 1).

**Figure 1:**
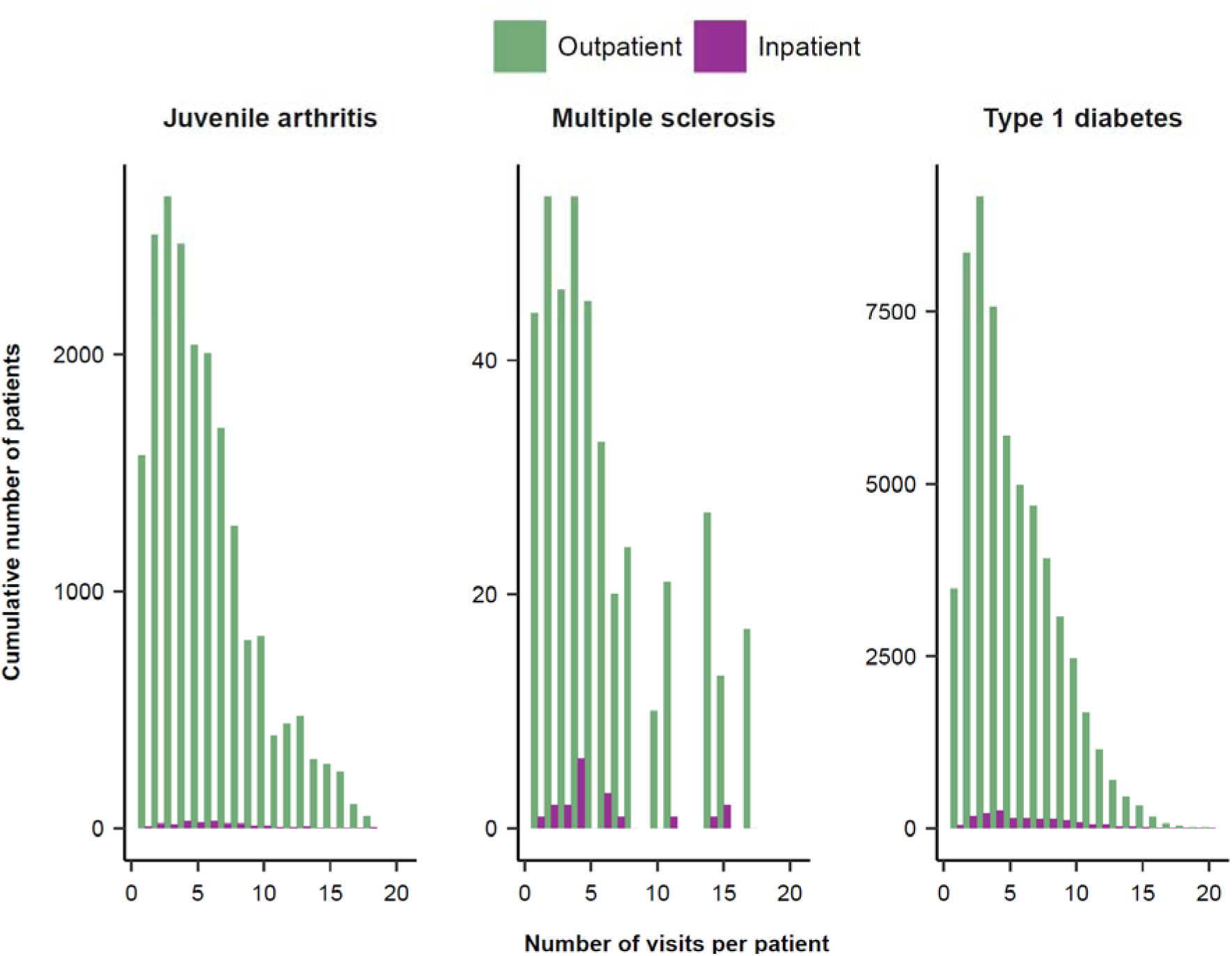
Number of patients with immune-mediated disease and number of hospital visits.

There was no consistent evidence of association between exposures and hospital visits in the 28-day and 180-day risk periods, except for a decreased relative risk for MS hospital visits after 3^rd^ dose (SCCS RR 0.33, 95%CI 0.15-0.71, cohort analysis RR 0.46, 95%CI 0.23-0.92, Table 4). There were some statistically significant results only observed with one analysis method. For T1D, there was an increased RR for T1D hospital visits after 2^nd^ dose in the 28-day risk period in the SCCS analysis (RR 1.09, 95%CI 1.04-1.14), and increased risk after the 1^st^ dose and 2^nd^ dose in the 180-days risk period in the SCCS analysis (RR 1,10, 95%CI 1.02-1.18, and RR 1.05, 95%CI 1.01-1.10, respectively). JRA had a higher relative risk in the 180-day study period after 1^st^ dose in the cohort analysis (RR 1.09, 95%CI 1.03-1.16) (Table 4).

**Table 4:**
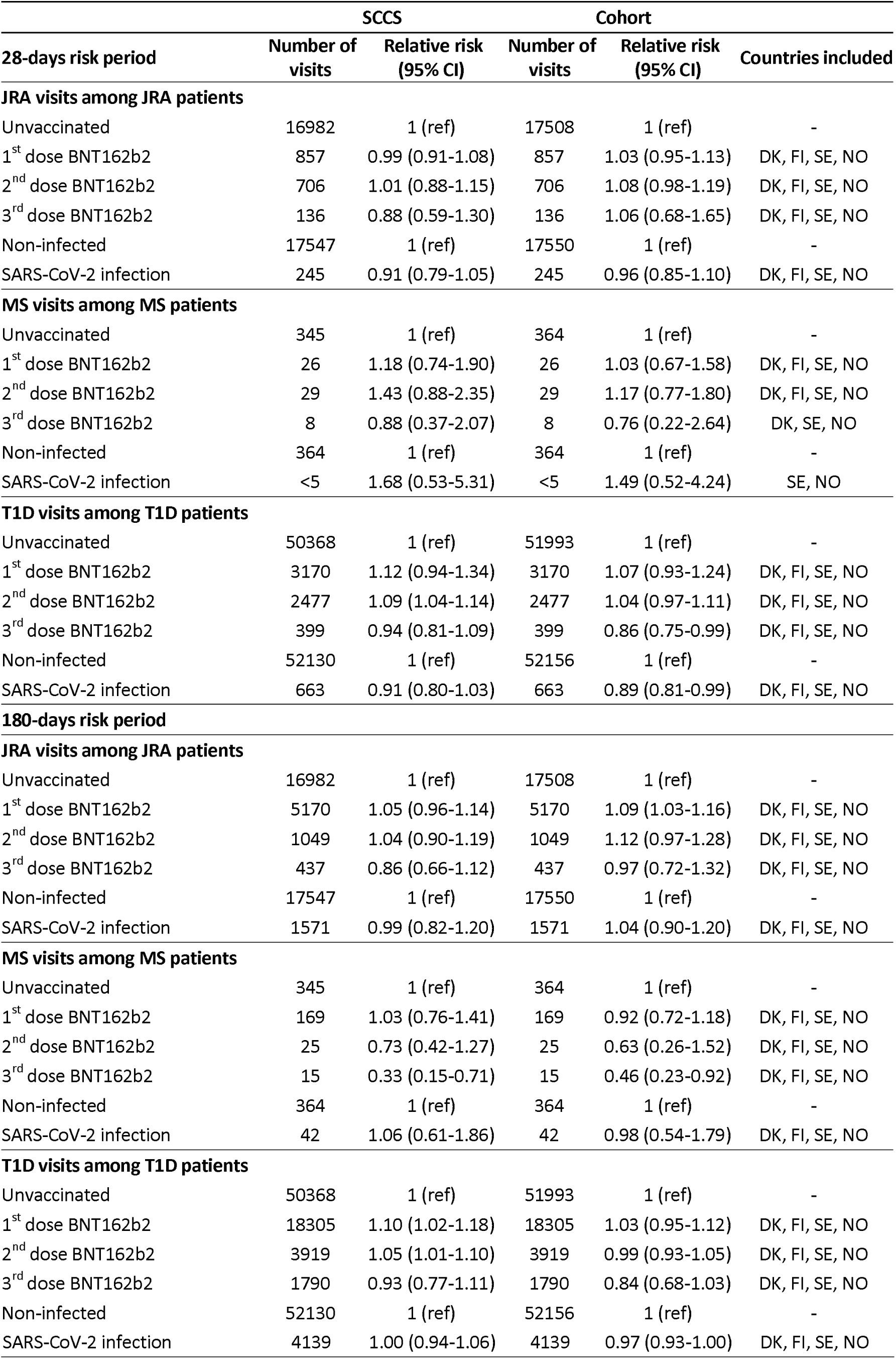
Relative risk of hospital visits.

|  | SCCS |  | Cohort |  |  |
| --- | --- | --- | --- | --- | --- |
| 28-days risk period | Number of visits | Relative risk (95% CI) | Number of visits | Relative risk (95% CI) | Countries included |
| JRA visits among JRA patients |  |  |  |  |  |
| Unvaccinated | 16982 | 1 (ref) | 17508 | 1 (ref) | - |
| 1<sup>st</sup> dose BNT162b2 | 857 | 0.99 (0.91-1.08) | 857 | 1.03 (0.95-1.13) | DK, FI, SE, NO |
| 2<sup>nd</sup> dose BNT162b2 | 706 | 1.01 (0.88-1.15) | 706 | 1.08 (0.98-1.19) | DK, FI, SE, NO |
| 3<sup>rd</sup> dose BNT162b2 | 136 | 0.88 (0.59-1.30) | 136 | 1.06 (0.68-1.65) | DK, FI, SE, NO |
| Non-infected | 17547 | 1 (ref) | 17550 | 1 (ref) | - |
| SARS-CoV-2 infection | 245 | 0.91 (0.79-1.05) | 245 | 0.96 (0.85-1.10) | DK, FI, SE, NO |
| MS visits among MS patients |  |  |  |  |  |
| Unvaccinated | 345 | 1 (ref) | 364 | 1 (ref) | - |
| 1<sup>st</sup> dose BNT162b2 | 26 | 1.18 (0.74-1.90) | 26 | 1.03 (0.67-1.58) | DK, FI, SE, NO |
| 2<sup>nd</sup> dose BNT162b2 | 29 | 1.43 (0.88-2.35) | 29 | 1.17 (0.77-1.80) | DK, FI, SE, NO |
| 3<sup>rd</sup> dose BNT162b2 | 8 | 0.88 (0.37-2.07) | 8 | 0.76 (0.22-2.64) | DK, SE, NO |
| Non-infected | 364 | 1 (ref) | 364 | 1 (ref) | - |
| SARS-CoV-2 infection | <5 | 1.68 (0.53-5.31) | <5 | 1.49 (0.52-4.24) | SE, NO |
| T1D visits among T1D patients |  |  |  |  |  |
| Unvaccinated | 50368 | 1 (ref) | 51993 | 1 (ref) | - |
| 1<sup>st</sup> dose BNT162b2 | 3170 | 1.12 (0.94-1.34) | 3170 | 1.07 (0.93-1.24) | DK, FI, SE, NO |
| 2<sup>nd</sup> dose BNT162b2 | 2477 | 1.09 (1.04-1.14) | 2477 | 1.04 (0.97-1.11) | DK, FI, SE, NO |
| 3<sup>rd</sup> dose BNT162b2 | 399 | 0.94 (0.81-1.09) | 399 | 0.86 (0.75-0.99) | DK, FI, SE, NO |
| Non-infected | 52130 | 1 (ref) | 52156 | 1 (ref) | - |
| SARS-CoV-2 infection | 663 | 0.91 (0.80-1.03) | 663 | 0.89 (0.81-0.99) | DK, FI, SE, NO |
| 180-days risk period |  |  |  |  |  |
| JRA visits among JRA patients |  |  |  |  |  |
| Unvaccinated | 16982 | 1 (ref) | 17508 | 1 (ref) | - |
| 1<sup>st</sup> dose BNT162b2 | 5170 | 1.05 (0.96-1.14) | 5170 | 1.09 (1.03-1.16) | DK, FI, SE, NO |
| 2<sup>nd</sup> dose BNT162b2 | 1049 | 1.04 (0.90-1.19) | 1049 | 1.12 (0.97-1.28) | DK, FI, SE, NO |
| 3<sup>rd</sup> dose BNT162b2 | 437 | 0.86 (0.66-1.12) | 437 | 0.97 (0.72-1.32) | DK, FI, SE, NO |
| Non-infected | 17547 | 1 (ref) | 17550 | 1 (ref) | - |
| SARS-CoV-2 infection | 1571 | 0.99 (0.82-1.20) | 1571 | 1.04 (0.90-1.20) | DK, FI, SE, NO |
| MS visits among MS patients |  |  |  |  |  |
| Unvaccinated | 345 | 1 (ref) | 364 | 1 (ref) | - |
| 1<sup>st</sup> dose BNT162b2 | 169 | 1.03 (0.76-1.41) | 169 | 0.92 (0.72-1.18) | DK, FI, SE, NO |
| 2<sup>nd</sup> dose BNT162b2 | 25 | 0.73 (0.42-1.27) | 25 | 0.63 (0.26-1.52) | DK, FI, SE, NO |
| 3<sup>rd</sup> dose BNT162b2 | 15 | 0.33 (0.15-0.71) | 15 | 0.46 (0.23-0.92) | DK, FI, SE, NO |
| Non-infected | 364 | 1 (ref) | 364 | 1 (ref) | - |
| SARS-CoV-2 infection | 42 | 1.06 (0.61-1.86) | 42 | 0.98 (0.54-1.79) | DK, FI, SE, NO |
| T1D visits among T1D patients |  |  |  |  |  |
| Unvaccinated | 50368 | 1 (ref) | 51993 | 1 (ref) | - |
| 1<sup>st</sup> dose BNT162b2 | 18305 | 1.10 (1.02-1.18) | 18305 | 1.03 (0.95-1.12) | DK, FI, SE, NO |
| 2<sup>nd</sup> dose BNT162b2 | 3919 | 1.05 (1.01-1.10) | 3919 | 0.99 (0.93-1.05) | DK, FI, SE, NO |
| 3<sup>rd</sup> dose BNT162b2 | 1790 | 0.93 (0.77-1.11) | 1790 | 0.84 (0.68-1.03) | DK, FI, SE, NO |
| Non-infected | 52130 | 1 (ref) | 52156 | 1 (ref) | - |
| SARS-CoV-2 infection | 4139 | 1.00 (0.94-1.06) | 4139 | 0.97 (0.93-1.00) | DK, FI, SE, NO |

In secondary analyses stratifying by age-groups and sex, there was a higher risk of MS hospital visits in boys during the 28-day risk period, and higher relative risks of hospital visits for T1D in the older age-groups (12-15 and 16-19-year-olds) in general (Supplemental Table 4).

## DISCUSSION

### Main findings

In this large (n=5,029,084), multinational Nordic cohort study of children and adolescents, we did not observe any robust support for associations between BNT162b2 vaccination and development of AIH, GBS or T1D, either in the 28-days or 180-days risk periods. We observed an association between SARS-CoV-2 infection and GBS.

### Strengths and limitations of the study

Although the present study is large, most outcomes are rare, which led to a lack of statistical precision in several estimates. The meta-analysis approach does not include estimates from a country if there were no outcomes in the unvaccinated or risk period, which could lead to overestimation of RRs. Results where not all countries contributed, such as the 3^rd^ dose BNT162b2-GBS association where only Norway contributed, should therefore be interpreted very carefully. Secondary analyses stratified by age-groups and sex are particularly susceptible to suffer from low number of cases. Like all observational studies, there will be unmeasured or residual confounding, particularly confounding by indication (e.g underlying health conditions associated with the outcomes are also associated with vaccination uptake or infection risk, giving rise to spurious associations). This is especially relevant in those receiving the 3^rd^ dose BNT162b2 at young ages. Although the SCCS method in theory accounts for all time-invariant confounding, thus avoiding potential confounding present in the cohort analysis, it is built on the assumption that outcomes do not influence future risk of exposure. This is likely to be violated in practice, especially in severe outcomes. The validity of hospital-register data is considered high,^25^ but there could be a diagnostic lag and there are likely erroneous registrations present in the registries. For diseases with a long development, such as T1D, any association during the 28-day risk period is unlikely to be causal and rather represents unmasking of existing disease, and the 180-day risk period could also be too brief to identify new-onset cases.^26^ Most participants do not have follow-up past 12 months and immune-mediated diseases could manifest beyond that. E.g. in a study of the 2009 pandemic swine influenza, the mean time from infection to diabetes diagnosis was two years.^3^ Infections, particularly asymptomatic infections, might have been unregistered, but there was extensive testing during the study period. As most hospital visits were outpatient visits, they could likely be “control” or routine visits, and potential increased disease activity signal could be hidden amongst routine visits. Vaccination could lead to more visits simply due to vaccination being recommended or mandatory prior to a planned control visit. A limitation is that the recommendation for spacing of vaccine doses might lead to lack of follow-up time in the 180-day risk period, particularly after the 1^st^ dose.

A strength of the study is the nationwide data from four Nordic countries with healthcare nominally free of charge for all residents, reducing selection bias. Using a CDM which harmonizes data structure and analysis across countries. Pooling results from different countries reduces the chance of country differences leading to spurious results, allows publishing of rare outcomes where each country might individually have too few cases, and results in one large study rather than four smaller studies in the literature. Another strength is the use of two different statistical methods that complement each other, with the SCCS controlling for time-invariant confounding. A common limitation in SARS-CoV-2 vaccine studies is that vaccine uptake is very rapid and high, leading to no contemporary reference group, which could be problematic if there are strong time trends. As the vaccination push was not so strong for children and adolescents, and by combining four countries we mitigate potential temporal effects as vaccination waves differed between countries. We had access to what we consider the strongest potential confounders and could adjust for these in the cohort analysis. Limiting outcomes to hospital contacts mitigate any potential vigilance bias, although it cannot completely be ruled out.

### Comparison with other studies

It is expected that some disease onset, or a flare-up of disease activity, would occur due to chance after vaccination, especially in a population-wide vaccination campaign setting. This will give rise to case reports, which are insufficient for establishing causality, but could warrant further studies.

There are unfortunately few earlier studies in children and adolescents for most outcomes studied. To our knowledge, this is the first cohort study reporting on new-onset AIH after BNT162b2 vaccination in children and adolescents. Lai et al. investigated a 28-day risk window after 1^st^ and 2^nd^ BNT162b2 vaccination in adolescents (12-18-year-olds, n=274,881), but found no GBS cases and only four T1D cases.^11^ Choe et al. found no GBS cases after BNT162b2 vaccination in adolescents (12-17-year-olds, n=2306) with chronic kidney disease.^27^ A prior study from the Norwegian investigators found too few GBS cases for a meaningful analysis in adolescents (12-19-year-olds, n=496,432), using Norwegian data,^28^ as did a recent English study (5-17-year-olds, n=5.1 million).^10^ Kim et al. report less GBS in a disproportionality analysis comparing COVID-19 vaccines vs other vaccines in adolescents (12-17 years), which is difficult to compare with our results.^29^ Hu et al. found no safety signal for GBS in children (5-17-year-olds, n= 3,017,352).^30^ A recent review (with mainly adult case-reports) report that GBS following COVID-19 vaccination seems to be more common after DNA vaccines and in older individuals, than after mRNA vaccines and younger individuals.^31^ The common theme in studies in children and adolescents seems to be that GBS is rare after SARS-CoV-2 vaccination, which fits our results and underlines the need for international collaboration to study rare events. It is uncertain whether there is increased risk of GBS after SARS-CoV-2 infection in adults,^32,33^ and there is a study presenting a high frequency (60%) of a preceding SARS-CoV-2 infection in children with GBS.^34^ Kildegaard et al. have too few GBS cases after SARS-CoV-2 infection for analysis in a study of Danish children (<18 years, n= 991,682),^4^ as did a recent English study (5-17-year-olds, n=5.1 million).^10^ We observe increased risk, but few cases and the fact that only two countries were included in the meta-analysis means that our results should be interpreted very cautiously. Earlier works on vaccination and T1D do not show any significant association after vaccination.^11,35^ Studies concerning new-onset T1D after SARS-CoV-2 infection show conflicting results (reviewed in ^36,19^), with both no association,^35,37^ and increased risk.^38^

We found no statistically significant associations between vaccination, SARS-CoV-2 infection and increased frequency of hospital visits in any of the patient cohorts (JRA, MS and T1D). Earlier studies show similar results.^39–43^

### Interpretation/Conclusions

The current study provides evidence supporting that BNT162b2 vaccination is not associated with new-onset AIH, GBS or T1D, or recurrent hospital visits of JRA, GBS or T1D in children and adolescents. The association between SARS-CoV-2 infection and new-onset GBS should be kept in mind for future waves of SARS-CoV-2 and ideally be replicated in other populations.

Acknowledgement

The study was commissioned and funded by the European Medicines Agency (EMA). The protocol was pre-registered in the European Union electronic Register of Post-Authorisation Studies (EUPAS), and the submitted study report is available at https://catalogues.ema.europa.eu/node/3529/administrative-details. This manuscript expresses the authors’ opinion and may not be understood or quoted as being made on behalf of or reflecting the position of the EMA or one of its committees or working parties.

## Data availability

The authors do not have the opportunity to share individual-level data, due to the nature of the data sources used. Data are accessible for researchers pending ethical approval and successful application to data sources in each of the countries involved.

## Author contributions

Hviid and Thiesson had full access to the data in the study and take responsibility for the integrity of the data and the accuracy of the analysis.

Concept and design: Karlstad, Thiesson, Nieminen, Pihlström, Tapia, Gulseth, Bakken, Hviid, Ljung.

Acquisition, analysis, or interpretation of data: Karlstad, Thiesson, Nieminen, Pihlström, Tapia, Gulseth, Bakken, Hviid, Ljung.

Drafting of the manuscript: Tapia, Hviid.

Critical revision of the manuscript for important intellectual content: Karlstad, Thiesson, Nieminen, Pihlström, Tapia, Gulseth, Bakken, Nohynek, Hviid, Ljung.

Statistical analysis: Thiesson, Nieminen, Pihlström, Tapia, Bakken.

Obtained funding: Karlstad, Gulseth, Nohynek, Hviid, Ljung.

Supervision: Tapia, Karlstad, Hviid, Ljung.

## References

1. Ascherio A, Zhang SM, Hernan MA, et al. Hepatitis B vaccination and the risk of multiple sclerosis. N Engl J Med. Feb 1 2001;344(5):327–32. doi:10.1056/NEJM200102013440502

2. Conti F, Rezai S, Valesini G. Vaccination and autoimmune rheumatic diseases. Autoimmun Rev. Dec 2008;8(2):124–8. doi:10.1016/j.autrev.2008.07.007

3. Ruiz PLD, Tapia G, Bakken IJ, et al. Pandemic influenza and subsequent risk of type 1 diabetes: a nationwide cohort study. Diabetologia. Sep 2018;61(9):1996–2004. doi:10.1007/s00125-018-4662-7

4. Kildegaard H, Lund LC, Hojlund M, Stensballe LG, Pottegard A. Risk of adverse events after covid-19 in Danish children and adolescents and effectiveness of BNT162b2 in adolescents: cohort study. BMJ. Apr 11 2022;377:e068898. doi:10.1136/bmj-2021-068898

5. Størdal K, Ruiz PL, Greve-Isdahl M, et al. Risk factors for SARS-CoV-2 infection and hospitalisation in children and adolescents in Norway: a nationwide population-based study. BMJ Open. Mar 11 2022;12(3):e056549. doi:10.1136/bmjopen-2021-056549

6. Ali K, Berman G, Zhou H, et al. Evaluation of mRNA-1273 SARS-CoV-2 Vaccine in Adolescents. N Engl J Med. Dec 9 2021;385(24):2241–2251. doi:10.1056/NEJMoa2109522

7. Frenck RW, Jr., Klein NP, Kitchin N, et al. Safety, Immunogenicity, and Efficacy of the BNT162b2 Covid-19 Vaccine in Adolescents. N Engl J Med. Jul 15 2021;385(3):239–250. doi:10.1056/NEJMoa2107456

8. Creech CB, Anderson E, Berthaud V, et al. Evaluation of mRNA-1273 Covid-19 Vaccine in Children 6 to 11 Years of Age. N Engl J Med. May 26 2022;386(21):2011–2023. doi:10.1056/NEJMoa2203315

9. Walter EB, Talaat KR, Sabharwal C, et al. Evaluation of the BNT162b2 Covid-19 Vaccine in Children 5 to 11 Years of Age. N Engl J Med. Jan 6 2022;386(1):35–46. doi:10.1056/NEJMoa2116298

10. Copland E, Patone M, Saatci D, et al. Safety outcomes following COVID-19 vaccination and infection in 5.1 million children in England. Nat Commun. May 27 2024;15(1):3822. doi:10.1038/s41467-024-47745-z

11. Lai FTT, Chua GT, Chan EWW, et al. Adverse events of special interest following the use of BNT162b2 in adolescents: a population-based retrospective cohort study. Emerg Microbes Infect. Dec 2022;11(1):885–893. doi:10.1080/22221751.2022.2050952

12. Nordstrom P, Ballin M, Nordstrom A. Safety and effectiveness of monovalent COVID-19 mRNA vaccination and risk factors for hospitalisation caused by the omicron variant in 0.8 million adolescents: A nationwide cohort study in Sweden. PLoS Med. Feb 2023;20(2):e1004127. doi:10.1371/journal.pmed.1004127

13. Luxenburger H, Thimme R. SARS-CoV-2 and the liver: clinical and immunological features in chronic liver disease. Gut. Sep 2023;72(9):1783–1794. doi:10.1136/gutjnl-2023-329623

14. Guo M, Liu X, Chen X, Li Q. Insights into new-onset autoimmune diseases after COVID-19 vaccination. Autoimmun Rev. Jul 2023;22(7):103340. doi:10.1016/j.autrev.2023.103340

15. Zheng H, Zhang T, Xu Y, Lu X, Sang X. Autoimmune hepatitis after COVID-19 vaccination. Front Immunol. 2022;13:1035073. doi:10.3389/fimmu.2022.1035073

16. ElSawi HA, Elborollosy A. Immune-mediated adverse events post-COVID vaccination and types of vaccines: a systematic review and meta-analysis. Egypt J Intern Med. 2022;34(1):44. doi:10.1186/s43162-022-00129-5

17. Sah BK, Fatima Z, Sah RK, et al. Guillain-Barre syndrome following COVID-19 vaccination: a study of 70 case reports. Ann Med Surg (Lond). Apr 2024;86(4):2067–2080. doi:10.1097/MS9.0000000000001915

18. Censi S, Bisaccia G, Gallina S, Tomassini V, Uncini A. Guillain-Barre syndrome and COVID-19 vaccination: a systematic review and meta-analysis. J Neurol. Mar 2024;271(3):1063–1071. doi:10.1007/s00415-024-12186-7

19. Wong R, Lam E, Bramante CT, et al. Does COVID-19 Infection Increase the Risk of Diabetes? Current Evidence. Curr Diab Rep. Aug 2023;23(8):207–216. doi:10.1007/s11892-023-01515-1

20. Farisogullari B, Pinto AS, Machado PM. COVID-19-associated arthritis: an emerging new entity? RMD Open. Sep 2022;8(2)doi:10.1136/rmdopen-2021-002026

21. Tavazzi E, Pichiecchio A, Colombo E, et al. The Potential Role of SARS-CoV-2 Infection and Vaccines in Multiple Sclerosis Onset and Reactivation: A Case Series and Literature Review. Viruses. Jul 18 2023;15(7)doi:10.3390/v15071569

22. Karlstad O, Hovi P, Husby A, et al. SARS-CoV-2 Vaccination and Myocarditis in a Nordic Cohort Study of 23 Million Residents. JAMA Cardiol. Jun 1 2022;7(6):600–612. doi:10.1001/jamacardio.2022.0583

23. Whitaker HJ, Farrington CP, Spiessens B, Musonda P. Tutorial in biostatistics: the self-controlled case series method. Stat Med. May 30 2006;25(10):1768–97. doi:10.1002/sim.2302

24. Whitaker HJ, Hocine MN, Farrington CP. The methodology of self-controlled case series studies. Stat Methods Med Res. Feb 2009;18(1):7–26. doi:10.1177/0962280208092342

25. Nielsen GL, Sorensen HT, Pedersen AB, Sabroe S. Analyses of data quality in registries concerning diabetes mellitus--a comparison between a population based hospital discharge and an insulin prescription registry. J Med Syst. Feb 1996;20(1):1–10. doi:10.1007/BF02260869

26. Steck AK, Vehik K, Bonifacio E, et al. Predictors of Progression From the Appearance of Islet Autoantibodies to Early Childhood Diabetes: The Environmental Determinants of Diabetes in the Young (TEDDY). Diabetes Care. May 2015;38(5):808–13. doi:10.2337/dc14-2426

27. Choe YJ, Ahn YH, Gwak E, Jo E, Kim J, Choe SA. Safety of BNT162b2 mRNA COVID-19 vaccine in children with chronic kidney disease: a national population study from South Korea. Pediatr Nephrol. Feb 2024;39(2):625–629. doi:10.1007/s00467-023-06195-3

28. Larsen VB, Gunnes N, Gran JM, et al. Adverse Events Following SARS-CoV-2 mRNA Vaccination in Adolescents: A Norwegian Nationwide Register-Based Study. medRxiv. 2023:2023.12.13.23299926. doi:10.1101/2023.12.13.23299926

29. Kim DH, Kim JH, Oh IS, Choe YJ, Choe SA, Shin JY. Adverse Events Following COVID-19 Vaccination in Adolescents: Insights From Pharmacovigilance Study of VigiBase. J Korean Med Sci. Mar 4 2024;39(8):e76. doi:10.3346/jkms.2024.39.e76

30. Hu M, Wong HL, Feng Y, et al. Safety of the BNT162b2 COVID-19 Vaccine in Children Aged 5 to 17 Years. JAMA Pediatr. Jul 1 2023;177(7):710–717. doi:10.1001/jamapediatrics.2023.1440

31. Yu M, Nie S, Qiao Y, Ma Y. Guillain-Barre syndrome following COVID-19 vaccines: A review of literature. Front Immunol. 2023;14:1078197. doi:10.3389/fimmu.2023.1078197

32. Censi S, Bisaccia G, Gallina S, Tomassini V, Uncini A. Guillain-Barre syndrome and SARS-CoV-2 infection: a systematic review and meta-analysis on a debated issue and evidence for the ‘Italian factor’. Eur J Neurol. Feb 2024;31(2):e16094. doi:10.1111/ene.16094

33. Bishara H, Arbel A, Barnett-Griness O, et al. Association Between Guillain-Barre Syndrome and COVID-19 Infection and Vaccination: A Population-Based Nested Case-Control Study. Neurology. Nov 14 2023;101(20):e2035–e2042. doi:10.1212/WNL.0000000000207900

34. Pourbakhtyaran E, Heidari M, Akbari MG, et al. Childhood Guillain-Barre syndrome in the SARS-CoV-2 era: Is there any causative relation? Clin Case Rep. Dec 2022;10(12):e6772. doi:10.1002/ccr3.6772

35. Stene LC, Ruiz PL-D, Ljung R, et al. Type 1 diabetes risk and severity after SARS-CoV-2 infection or vaccination. medRxiv. 2024:2024.07.03.24309894. doi:10.1101/2024.07.03.24309894

36. Karavanaki K, Rodolaki K, Soldatou A, Karanasios S, Kakleas K. Covid-19 infection in children and adolescents and its association with type 1 diabetes mellitus (T1d) presentation and management. Endocrine. May 2023;80(2):237–252. doi:10.1007/s12020-022-03266-7

37. Noorzae R, Junker TG, Hviid AP, Wohlfahrt J, Olsen SF. Risk of Type 1 Diabetes in Children Is Not Increased After SARS-CoV-2 Infection: A Nationwide Prospective Study in Denmark. Diabetes Care. Jun 1 2023;46(6):1261–1264. doi:10.2337/dc22-2351

38. Barrett CE, Koyama AK, Alvarez P, et al. Risk for Newly Diagnosed Diabetes >30 Days After SARS-CoV-2 Infection Among Persons Aged <18 Years - United States, March 1, 2020-June 28, 2021. MMWR Morb Mortal Wkly Rep. Jan 14 2022;71(2):59–65. doi:10.15585/mmwr.mm7102e2

39. Blanco Y, Escudero D, Lleixa C, et al. mRNA COVID-19 Vaccination Does Not Exacerbate Symptoms or Trigger Neural Antibody Responses in Multiple Sclerosis. Neurol Neuroimmunol Neuroinflamm. Nov 2023;10(6)doi:10.1212/NXI.0000000000200163

40. van Dam KPJ, Wieske L, Stalman EW, et al. Disease activity in patients with immune-mediated inflammatory diseases after SARS-CoV-2 vaccinations. J Autoimmun. Feb 2023;135:102984. doi:10.1016/j.jaut.2022.102984

41. Opoka-Winiarska V, Lipinska J, Michalak A, Burzynski J, Kadziolka O, Smolewska E. Safety of the COVID-19 vaccination in children with juvenile idiopathic arthritis-A observational study from two pediatric rheumatology centres in Poland. Front Pediatr. 2023;11:1103763. doi:10.3389/fped.2023.1103763

42. Hamad Saied M, van Straalen JW, de Roock S, et al. Humoral and cellular immunogenicity, effectiveness and safety of COVID-19 mRNA vaccination in patients with pediatric rheumatic diseases: A prospective cohort study. Vaccine. Feb 15 2024;42(5):1145–1153. doi:10.1016/j.vaccine.2024.01.047

43. Achiron A, Dolev M, Menascu S, et al. COVID-19 vaccination in patients with multiple sclerosis: What we have learnt by February 2021. Mult Scler. May 2021;27(6):864–870. doi:10.1177/13524585211003476

