## supplemental material for "Association between Covid-19 BNT162b2 vaccination or SARS-CoV-2 infection and immune mediated diseases in 5 million 5-to-19-year-olds in four Nordic countries"

**Supplemental Methods**

### Ethics

*Denmark:* The Danish study was conducted using administrative register data, which according to Danish law does not require external ethics approval.

*Finland:* The Finnish study is a part of vaccine surveillance work, which is one of the duties of the Finnish Institute for Health and Welfare (THL). For this work, the institute is obliged to utilize all available data, including register data, to investigate potential harmful effects of the vaccines. Consent to participate was not applicable as this is a register-based study.

*Norway:* The Norwegian study was approved by the Norwegian Regional Committee for Health Research Ethics South-East (REK Sør-Øst A, ref 122745). The emergency preparedness register BeredtC19 which provided data was established according to the Health Preparedness Act §2-4. Consent to participate was not applicable as this is a register-based study.

*Sweden:* The Swedish study is approved by the Swedish Ethical Review Authority (2020-06859, 2021-02186). Consent to participate is not applicable as this is a register-based study.

**Supplementary Table 1: Outcomes, comorbities and ICD-10 codes used.**

| **Study outcome** | **ICD-10 codes used to identify cases** | |
| --- | --- | --- |
|  | ICD-10 codes with exactly 3-digits | ICD-10 codes with 4-digits |
| Autoimmune hepatitis |  | K754 |
| Juvenile rheumatoid arthritis | M08 | M080, M082, M083, M084, -M088, M089 |
| Guillain-Barré syndrome |  | G610 |
| Multiple sclerosis | G35 | G359 |
| Type 1 diabetes | E10 | E100, E101, E102, E103, E104, E105, E106, E107, E108, E109 |
| **Covariates (registered prior to first covid vaccination)** |  |  |
| Asthma | J45-46 |  |
| Other chronic respiratory diseases | E84, J41-44, J47, J84, P27 |  |
| Chronic cardiac disease | I05-08, I20-28, I34-37, I42-49, I50-51 |  |
| Renal disease | N03, N05, N07, N18, N19, N25-27 |  |
| Epilepsy or convulsions | G40, R56 |  |
| Congenital malformations and chromosomal abnormalities | Q00-07, Q20-28, Q30-34, Q60-64, Q90-99 |  |
| Malignancy or immunodeficiency | C00-96, D70-72, D81-84 | D730 |
| Psychiatric disorder | Any chapter F diagnosis |  |

**Supplemental Table 2: sensitivity analyses of new-onset disease.**

| **28 days** | **SCCS** |  |  |  |  | **Cohort** | | | | |
| --- | --- | --- | --- | --- | --- | --- | --- | --- | --- | --- |
| **Exposure** | **Boys** | **Girls** | **Age 5 to 11** | **Age 12 to 15** | **Age 16 to 19** | **Boys** | **Girls** | **Age 5 to 11** | **Age 12 to 15** | **Age 16 to 19** |
| **Autoimmune hepatitis** |  |  |  |  |  |  |  |  |  |  |
| Unvaccinated | 1 (ref) | 1 (ref) | 1 (ref) | 1 (ref) | 1 (ref) | 1 (ref) | 1 (ref) | 1 (ref) | 1 (ref) | 1 (ref) |
| 1^st^ dose BNT162b2 | NA | NA | NA | NA | NA | NA | NA | NA | NA | NA |
| 2^nd^ dose BNT162b2 | NA | 1.45 (0.10-20.83) | NA | NA | 0.44 (0.02-9.49) | NA | 1.98 (0.21-18.88) | NA | NA | 1.19 (0.13-10.46) |
| 3^rd^ dose BNT1262b2 | NA | NA | NA | NA | NA | NA | NA | NA | NA | NA |
| Non-infected | 1 (ref) | 1 (ref) | 1 (ref) | 1 (ref) | 1 (ref) | 1 (ref) | 1 (ref) | 1 (ref) | 1 (ref) | 1 (ref) |
| SARS-CoV-2 infection | NA | NA | NA | NA | 2.19 (0.09-52.82) | 14.25 (1.73-117.51) | NA | NA | NA | 10.97 (1.13-106.22) |
| **Guillain-Barré syndrome** |  |  |  |  |  |  |  |  |  |  |
| Unvaccinated | 1 (ref) | 1 (ref) | 1 (ref) | 1 (ref) | 1 (ref) | 1 (ref) | 1 (ref) | 1 (ref) | 1 (ref) | 1 (ref) |
| 1^st^ dose BNT162b2 | NA | NA | NA | NA | NA | NA | NA | NA | NA | NA |
| 2^nd^ dose BNT162b2 | NA | NA | NA | NA | NA | NA | NA | NA | NA | NA |
| 3^rd^ dose BNT1262b2 | NA | NA | NA | NA | NA | NA | NA | NA | NA | NA |
| Non-infected | 1 (ref) | 1 (ref) | 1 (ref) | 1 (ref) | 1 (ref) | 1 (ref) | 1 (ref) | 1 (ref) | 1 (ref) | 1 (ref) |
| SARS-CoV-2 infection | NA | NA | 48.53 (2.60-905.30) | NA | NA | 3.28 (0.27-40.11) | 57.37 (8.93-368.40) | 33.69 (5.57-203.90) | NA | 18.29 (0.78-430.15) |
| **Type 1 diabetes** |  |  |  |  |  |  |  |  |  |  |
| Unvaccinated | 1 (ref) | 1 (ref) | 1 (ref) | 1 (ref) | 1 (ref) | 1 (ref) | 1 (ref) | 1 (ref) | 1 (ref) | 1 (ref) |
| 1^st^ dose BNT162b2 | 0.98 (0.73-1.32) | 0.58 (0.31-1.08) | 0.55 (0.17-1.74) | 0.94 (0.61-1.44) | 1.26 (0.81-1.97) | 1.32 (0.99-1.75) | 0.84 (0.54-1.29) | 1.35 (0.29-6.35) | 1.23 (0.88-1.73) | 1.42 (0.94-2.14) |
| 2^nd^ dose BNT162b2 | 0.74 (0.50-1.12) | 0.57 (0.35-0.93) | 0.34 (0.10-1.11) | 0.58 (0.34-0.98) | 1.16 (0.69-1.94) | 1.12 (0.72-1.75) | 0.91 (0.57-1.46) | 0.92 (0.29-2.89) | 0.91 (0.56-1.48) | 1.52 (0.97-2.39) |
| 3^rd^ dose BNT1262b2 | 0.97 (0.12-7.67) | 2.55 (0.25-26.20) | NA | NA | 0.83 (0.17-3.97) | 2.14 (0.29-15.65) | 2.43 (0.31-18.87) | NA | NA | 1.03 (0.24-4.46) |
| Non-infected | 1 (ref) | 1 (ref) | 1 (ref) | 1 (ref) | 1 (ref) | 1 (ref) | 1 (ref) | 1 (ref) | 1 (ref) | 1 (ref) |
| SARS-CoV-2 infection | 1.21 (0.71-2.06) | 1.42 (0.52-3.89) | 1.05 (0.57-1.92) | 1.47 (0.66-3.29) | 3.78 (1.43-9.99) | 1.36 (0.83-2.20) | 1.25 (0.47-3.30) | 1.07 (0.66-1.72) | 1.50 (0.87-2.58) | 2.74 (1.33-5.63) |
| **180 days** |  |  |  |  |  |  |  |  |  |  |
| **Autoimmune hepatitis** |  |  |  |  |  |  |  |  |  |  |
| Unvaccinated | 1 (ref) | 1 (ref) | 1 (ref) | 1 (ref) | 1 (ref) | 1 (ref) | 1 (ref) | 1 (ref) | 1 (ref) | 1 (ref) |
| 1^st^ dose BNT162b2 | 0.36 (0.04-2.97) | 1.15 (0.21-6.13) | NA | 0.93 (0.07-11.92) | 0.39 (0.10-1.53) | NA | NA | NA | NA | NA |
| 2^nd^ dose BNT162b2 | 0.27 (0.01-5.19) | NA | NA | NA | 0.16 (0.01-4.32) | NA | 1.98 (0.21-18.88) | NA | NA | 1.19 (0.13-10.46) |
| 3^rd^ dose BNT1262b2 | NA | 3.37 (0.10-118.05) | NA | NA | 0.44 (0.02-8.03) | NA | NA | NA | NA | NA |
| Non-infected | 1 (ref) | 1 (ref) | 1 (ref) | 1 (ref) | 1 (ref) | 1 (ref) | 1 (ref) | 1 (ref) | 1 (ref) | 1 (ref) |
| SARS-CoV-2 infection | NA | 0.55 (0.03-8.94) | NA | NA | 0.93 (0.02-37.38) | 14.25 (1.73-117.51) | NA | NA | NA | 10.97 (1.13-106.22) |
| **Guillain-Barré syndrome** |  |  |  |  |  |  |  |  |  |  |
| Unvaccinated | 1 (ref) | 1 (ref) | 1 (ref) | 1 (ref) | 1 (ref) | 1 (ref) | 1 (ref) | 1 (ref) | 1 (ref) | 1 (ref) |
| 1^st^ dose BNT162b2 | 0.87 (0.10-7.72) | NA | NA | 8.15 (0.05-1251.28) | NA | 1.00 (0.20-5.00) | 13.23 (0.12-1416.23) | NA | 2.13 (0.06-80.82) | 11.42 (0.14-924.50) |
| 2^nd^ dose BNT162b2 | NA | NA | NA | NA | NA | NA | NA | NA | NA | NA |
| 3^rd^ dose BNT1262b2 | NA | NA | NA | NA | NA | NA | NA | NA | NA | NA |
| Non-infected | 1 (ref) | 1 (ref) | 1 (ref) | 1 (ref) | 1 (ref) | 1 (ref) | 1 (ref) | 1 (ref) | 1 (ref) | 1 (ref) |
| SARS-CoV-2 infection | NA | NA | 38.76 (1.33-1131.97) | NA | NA | 2.03 (0.27-15.53) | 7.96 (2.28-27.79) | 4.47 (1.34-14.90) | 6.64 (0.68-64.96) | 2.91 (0.15-56.32) |
| **Type 1 diabetes** |  |  |  |  |  |  |  |  |  |  |
| Unvaccinated | 1 (ref) | 1 (ref) | 1 (ref) | 1 (ref) | 1 (ref) | 1 (ref) | 1 (ref) | 1 (ref) | 1 (ref) | 1 (ref) |
| 1^st^ dose BNT162b2 | 0.83 (0.64-1.06) | 0.63 (0.47-0.84) | 0.51 (0.36-0.73) | 0.74 (0.57-0.98) | 1.09 (0.77-1.53) | 1.24 (1.03-1.49) | 0.96 (0.78-1.18) | 1.23 (0.92-1.64) | 1.12 (0.88-1.42) | 1.30 (1.00-1.69) |
| 2^nd^ dose BNT162b2 | 0.49 (0.34-0.71) | 0.70 (0.36-1.35) | 0.49 (0.22-1.07) | 0.60 (0.36-1.01) | 0.91 (0.54-1.53) | 0.99 (0.70-1.40) | 1.34 (0.58-3.07) | 1.36 (0.65-2.82) | 1.06 (0.47-2.41) | 1.26 (0.83-1.93) |
| 3^rd^ dose BNT1262b2 | 0.42 (0.20-0.87) | 0.63 (0.29-1.39) | NA | 0.24 (0.03-2.05) | 0.79 (0.38-1.66) | 1.08 (0.56-2.07) | 1.78 (0.92-3.47) | NA | 1.04 (0.14-7.64) | 1.43 (0.80-2.55) |
| Non-infected | 1 (ref) | 1 (ref) | 1 (ref) | 1 (ref) | 1 (ref) | 1 (ref) | 1 (ref) | 1 (ref) | 1 (ref) | 1 (ref) |
| SARS-CoV-2 infection | 0.90 (0.62-1.28) | 0.91 (0.64-1.27) | 0.83 (0.52-1.33) | 1.04 (0.68-1.61) | 1.72 (0.82-3.59) | 1.03 (0.80-1.33) | 0.83 (0.64-1.07) | 0.85 (0.67-1.08) | 1.00 (0.75-1.32) | 1.16 (0.77-1.74) |

*NA: Not Available*

**Supplemental Table 3: Characteristics of the study cohorts of patients with pre-existing immune-mediated diseases.**

| **Characteristic** | **Unvaccinated N (%^1^)** | **1st dose** **N (%^1^)** | **2^nd^ dose N (%^1^)** | **3^rd^ dose N (%^1^)** | **Other N (%^1^)** | **Infected N (%^1^)** | **Non-infected N (%^1^)** | **Total** |
| --- | --- | --- | --- | --- | --- | --- | --- | --- |
| **Juvenile arthritis patient cohort** | | | | | | | | |
| **Denmark** | 450 (23.3%) | 70 (3.6%) | 1037 (53.8%) | 364 (18.9%) | 7 (0.4%) | 1474 (76.5%) | 454 (23.5%) | 1928 |
| **Finland** | 1001 (31.2%) | 221 (6.9%) | 1074 (33.4%) | 461 (14.4%) | 454 (14.1%) | 826 (25.7%) | 2385 (74.3%) | 3211 |
| **Norway** | 510 (35.7%) | 299 (20.9%) | 384 (26.9%) | 142 (9.9%) | 94 (6.6%) | 693 (48.5%) | 736 (51.5%) | 1429 |
| **Sweden** | 391 (18.5%) | 70 (3.3%) | 1084 (51.3%) | 315 (14.9%) | 252 (11.9%) | 670 (31.7%) | 1442 (68.3%) | 2112 |
| **Girls** | 1445 (26.6%) | 404 (7.4%) | 2175 (40.0%) | 859 (15.8%) | 553 (10.2%) | 2307 (42.4%) | 3129 (57.6%) | 5436 |
| **Boys** | 907 (28.0%) | 256 (7.9%) | 1404 (43.3%) | 423 (13.0%) | 254 (7.8%) | 1356 (41.8%) | 1888 (58.2%) | 3244 |
| **Age 5 to 11** | 1366 (65.5%) | 237 (11.4%) | 473 (22.7%) | 9 (0.4%) | <5 | 899 (43.1%) | 1187 (56.9%) | 2086 |
| **Age 12 to 15** | 616 (20.2%) | 314 (10.3%) | 1687 (55.3%) | 151 (5.0%) | 280 (9.2%) | 1267 (41.6%) | 1781 (58.4%) | 3048 |
| **Age 16 to 19** | 370 (10.4%) | 109 (3.1%) | 1419 (40.0%) | 1122 (31.6%) | 526 (14.8%) | 1497 (42.2%) | 2049 (57.8%) | 3546 |
| **0 comorbidity** | 1875 (27.6%) | 531 (7.8%) | 2829 (41.6%) | 949 (14.0%) | 613 (9.0%) | 2967 (43.7%) | 3830 (56.3%) | 6797 |
| **1 comorbidity** | 397 (25.1%) | 105 (6.6%) | 650 (41.1%) | 265 (16.8%) | 163 (10.3%) | 582 (36.8%) | 998 (63.2%) | 1580 |
| **2+ comorbidities** | 80 (26.4%) | 24 (7.9%) | 100 (33.0%) | 68 (22.4%) | 31 (10.2%) | 114 (37.6%) | 189 (62.4%) | 303 |
| **Total** | 2352 (27.1%) | 660 (7.6%) | 3579 (41.2%) | 1282 (14.8%) | 807 (9.3%) | 3663 (42.2%) | 5017 (57.8%) | 8680 |
| **Multiple sclerosis patient cohort** | | | | | | | | |
| **Denmark** | 7 (17.5%) | <5 | 8 (20.0%) | 24 (60.0%) | 0 (0.0%) | 31 (77.5%) | 9 (22.5%) | 40 |
| **Finland** | <5 | <5 | <5 | 7 (38.9%) | 5 (27.8%) | 10 (55.6%) | 8 (44.4%) | 18 |
| **Norway** | <5 | <5 | 19 (42.2%) | 10 (22.2%) | 8 (17.8%) | 24 (53.3%) | 21 (46.7%) | 45 |
| **Sweden** | 14 (23.7%) | <5 | 20 (33.9%) | 17 (28.8%) | 7 (11.9%) | 18 (30.5%) | 41 (69.5%) | 59 |
| **Girls** | 16 (14.3%) | <5 | 35 (31.2%) | 43 (38.4%) | 15 (13.4%) | 57 (50.9%) | 55 (49.1%) | 112 |
| **Boys** | 11 (22.0%) | <5 | 15 (30.0%) | 15 (30.0%) | 5 (10.0%) | 26 (52.0%) | 24 (48.0%) | 50 |
| **Age 5 to 11** | <5 | 0 (0.0%) | <5 | 0 (0.0%) | 0 (0.0%) | <5 | 0 (0.0%) | <5 |
| **Age 12 to 15** | 7 (20.6%) | 5 (14.7%) | 13 (38.2%) | 6 (17.6%) | <5 | 15 (44.1%) | 19 (55.9%) | 34 |
| **Age 16 to 19** | 19 (15.1%) | <5 | 36 (28.6%) | 52 (41.3%) | 17 (13.5%) | 66 (52.4%) | 60 (47.6%) | 126 |
| **0 comorbidity** | 15 (12.5%) | <5 | 42 (35.0%) | 48 (40.0%) | 12 (10.0%) | 59 (49.2%) | 61 (50.8%) | 120 |
| **1 comorbidity** | 10 (28.6%) | <5 | 7 (20.0%) | 9 (25.7%) | 7 (20.0%) | 21 (60.0%) | 14 (40.0%) | 35 |
| **2+ comorbidities** | <5 | <5 | <5 | <5 | <5 | <5 | <5 | 7 |
| **Total** | 27 (16.7%) | 7 (4.3%) | 50 (30.9%) | 58 (35.8%) | 20 (12.3%) | 83 (51.2%) | 79 (48.8%) | 162 |
| **Type 1 diabetes patient cohort** | | | | | | | | |
| **Denmark** | 557 (18.1%) | 85 (2.8%) | 1760 (57.1%) | 660 (21.4%) | 22 (0.7%) | 2194 (71.1%) | 890 (28.9%) | 3084 |
| **Finland** | 1486 (23.8%) | 477 (7.6%) | 2401 (38.4%) | 1190 (19.0%) | 698 (11.2%) | 1556 (24.9%) | 4696 (75.1%) | 6252 |
| **Norway** | 1294 (34.5%) | 906 (24.1%) | 924 (24.6%) | 402 (10.7%) | 229 (6.1%) | 1589 (42.3%) | 2166 (57.7%) | 3755 |
| **Sweden** | 895 (15.4%) | 195 (3.4%) | 3094 (53.2%) | 981 (16.9%) | 649 (11.2%) | 1650 (28.4%) | 4164 (71.6%) | 5814 |
| **Girls** | 1929 (22.2%) | 742 (8.5%) | 3730 (42.9%) | 1503 (17.3%) | 797 (9.2%) | 3376 (38.8%) | 5325 (61.2%) | 8701 |
| **Boys** | 2303 (22.6%) | 921 (9.0%) | 4449 (43.6%) | 1730 (17.0%) | 801 (7.8%) | 3613 (35.4%) | 6591 (64.6%) | 10204 |
| **Age 5 to 11** | 2252 (62.4%) | 463 (12.8%) | 871 (24.1%) | 19 (0.5%) | 6 (0.2%) | 1501 (41.6%) | 2110 (58.4%) | 3611 |
| **Age 12 to 15** | 1177 (17.3%) | 947 (13.9%) | 3979 (58.4%) | 315 (4.6%) | 399 (5.9%) | 2471 (36.2%) | 4346 (63.8%) | 6817 |
| **Age 16 to 19** | 803 (9.5%) | 253 (3.0%) | 3329 (39.3%) | 2899 (34.2%) | 1193 (14.1%) | 3017 (35.6%) | 5460 (64.4%) | 8477 |
| **0 comorbidity** | 3321 (22.7%) | 1339 (9.2%) | 6374 (43.6%) | 2406 (16.5%) | 1163 (8.0%) | 5583 (38.2%) | 9020 (61.8%) | 14603 |
| **1 comorbidity** | 772 (20.6%) | 278 (7.4%) | 1614 (43.0%) | 708 (18.9%) | 383 (10.2%) | 1233 (32.8%) | 2522 (67.2%) | 3755 |
| **2+ comorbidities** | 139 (25.4%) | 46 (8.4%) | 191 (34.9%) | 119 (21.8%) | 52 (9.5%) | 173 (31.6%) | 374 (68.4%) | 547 |
| **Total** | 4232 (22.4%) | 1663 (8.8%) | 8179 (43.3%) | 3233 (17.1%) | 1598 (8.5%) | 6989 (37.0%) | 11916 (63.0%) | 18905 |

^1^ Row percentages

**Supplemental Table 4: Hospital visits/flares stratified by sex and age**

|  | **SCCS** |  |  |  |  | **Contemporary cohort** | | | | |
| --- | --- | --- | --- | --- | --- | --- | --- | --- | --- | --- |
| **28 days** | **Boys** | **Girls** | **Age 5 to 11** | **Age 12 to 15** | **Age 16 to 19** | **Boys** | **Girls** | **Age 5 to 11** | **Age 12 to 15** | **Age 16 to 19** |
| **JRA** |  |  |  |  |  |  |  |  |  |  |
| Unvaccinated | 1 (ref) | 1 (ref) | 1 (ref) | 1 (ref) | 1 (ref) | 1 (ref) | 1 (ref) | 1 (ref) | 1 (ref) | 1 (ref) |
| 1^st^ dose BNT162b2 | 0.95 (0.83-1.09) | 1.00 (0.90-1.12) | 0.82 (0.61-1.10) | 1.11 (0.94-1.31) | 0.96 (0.85-1.09) | 1.02 (0.90-1.16) | 1.04 (0.93-1.16) | 0.91 (0.74-1.11) | 1.10 (0.97-1.25) | 1.01 (0.90-1.13) |
| 2^nd^ dose BNT162b2 | 0.98 (0.84-1.14) | 1.03 (0.89-1.19) | 0.86 (0.58-1.30) | 0.99 (0.71-1.39) | 1.09 (0.95-1.26) | 1.07 (0.87-1.32) | 1.08 (0.98-1.19) | 0.95 (0.69-1.31) | 1.03 (0.85-1.26) | 1.16 (1.02-1.31) |
| 3^rd^ dose BNT1262b2 | 0.84 (0.41-1.72) | 0.99 (0.80-1.24) | NA | 1.37 (0.93-2.03) | 1.05 (0.65-1.70) | 0.98 (0.42-2.27) | 1.20 (0.97-1.47) | NA | 1.58 (1.10-2.25) | 1.18 (0.69-2.00) |
| Non-infected | 1 (ref) | 1 (ref) | 1 (ref) | 1 (ref) | 1 (ref) | 1 (ref) | 1 (ref) | 1 (ref) | 1 (ref) | 1 (ref) |
| SARS-CoV-2 infection | 0.94 (0.74-1.18) | 0.90 (0.75-1.07) | 1.00 (0.82-1.23) | 0.79 (0.61-1.02) | 0.93 (0.68-1.26) | 1.03 (0.83-1.28) | 0.93 (0.79-1.09) | 1.04 (0.86-1.25) | 0.85 (0.68-1.08) | 0.94 (0.72-1.24) |
| **MS** |  |  |  |  |  |  |  |  |  |  |
| Unvaccinated | 1 (ref) | 1 (ref) | 1 (ref) | 1 (ref) | 1 (ref) | 1 (ref) | 1 (ref) | 1 (ref) | 1 (ref) | 1 (ref) |
| 1^st^ dose BNT162b2 | 3.17 (1.15-8.76) | 0.74 (0.40-1.35) | NA | 2.00 (0.69-5.82) | 1.08 (0.62-1.86) | 2.69 (1.08-6.69) | 0.67 (0.39-1.18) | NA | 1.62 (0.58-4.50) | 1.04 (0.64-1.71) |
| 2^nd^ dose BNT162b2 | 3.01 (0.88-10.24) | 1.16 (0.63-2.13) | NA | 17.57 (3.62-85.40) | 1.36 (0.69-2.65) | 2.66 (1.02-6.95) | 0.87 (0.50-1.51) | NA | 10.27 (2.42-43.54) | 1.21 (0.72-2.04) |
| 3^rd^ dose BNT1262b2 | 15.01 (1.78-126.82) | 0.72 (0.25-2.07) | NA | NA | 0.79 (0.32-1.97) | 22.93 (5.22-100.83) | 0.41 (0.16-1.08) | NA | NA | 0.74 (0.22-2.52) |
| Non-infected | 1 (ref) | 1 (ref) | 1 (ref) | 1 (ref) | 1 (ref) | 1 (ref) | 1 (ref) | 1 (ref) | 1 (ref) | 1 (ref) |
| SARS-CoV-2 infection | 1.22 (0.21-7.01) | 1.68 (0.34-8.38) | NA | 1.38 (0.11-16.95) | 0.86 (0.17-4.29) | 1.66 (0.37-7.41) | 1.56 (0.34-7.14) | NA | 2.12 (0.15-29.66) | 0.71 (0.17-2.96) |
| **Type 1 diabetes** |  |  |  |  |  |  |  |  |  |  |
| Unvaccinated | 1 (ref) | 1 (ref) | 1 (ref) | 1 (ref) | 1 (ref) | 1 (ref) | 1 (ref) | 1 (ref) | 1 (ref) | 1 (ref) |
| 1^st^ dose BNT162b2 | 1.13 (0.95-1.33) | 1.11 (0.91-1.36) | 0.90 (0.78-1.03) | 1.17 (0.84-1.63) | 1.22 (1.05-1.41) | 1.08 (0.93-1.25) | 1.06 (0.91-1.23) | 0.92 (0.81-1.04) | 1.12 (0.85-1.47) | 1.11 (1.00-1.24) |
| 2^nd^ dose BNT162b2 | 1.09 (1.02-1.16) | 1.09 (1.02-1.17) | 0.89 (0.74-1.07) | 1.20 (0.96-1.49) | 1.18 (1.05-1.32) | 1.03 (0.94-1.12) | 1.04 (0.98-1.12) | 0.93 (0.71-1.22) | 1.12 (0.88-1.42) | 1.05 (0.98-1.12) |
| 3^rd^ dose BNT1262b2 | 0.98 (0.79-1.22) | 0.92 (0.75-1.13) | NA | 1.25 (0.64-2.42) | 1.07 (0.94-1.22) | 0.88 (0.76-1.01) | 0.83 (0.65-1.05) | NA | 1.12 (0.66-1.89) | 0.81 (0.72-0.91) |
| Non-infected | 1 (ref) | 1 (ref) | 1 (ref) | 1 (ref) | 1 (ref) | 1 (ref) | 1 (ref) | 1 (ref) | 1 (ref) | 1 (ref) |
| SARS-CoV-2 infection | 0.88 (0.71-1.10) | 0.93 (0.83-1.06) | 0.85 (0.69-1.04) | 0.92 (0.79-1.08) | 1.01 (0.85-1.21) | 0.87 (0.72-1.05) | 0.91 (0.81-1.02) | 0.85 (0.74-0.98) | 0.93 (0.80-1.09) | 0.94 (0.81-1.09) |
| **180 days** |  |  |  |  |  |  |  |  |  |  |
| **JRA** |  |  |  |  |  |  |  |  |  |  |
| Unvaccinated | 1 (ref) | 1 (ref) | 1 (ref) | 1 (ref) | 1 (ref) | 1 (ref) | 1 (ref) | 1 (ref) | 1 (ref) | 1 (ref) |
| 1^st^ dose BNT162b2 | 1.03 (0.93-1.14) | 1.05 (0.95-1.18) | 0.93 (0.73-1.18) | 1.12 (0.93-1.36) | 1.08 (0.99-1.19) | 1.10 (0.95-1.27) | 1.10 (1.04-1.16) | 1.04 (0.90-1.20) | 1.08 (0.97-1.20) | 1.14 (1.06-1.22) |
| 2^nd^ dose BNT162b2 | 1.02 (0.87-1.20) | 1.05 (0.91-1.23) | 1.03 (0.76-1.39) | 1.17 (0.92-1.48) | 1.18 (0.97-1.44) | 1.14 (0.85-1.52) | 1.12 (1.03-1.22) | 1.18 (0.93-1.49) | 1.16 (0.93-1.45) | 1.24 (1.04-1.47) |
| 3^rd^ dose BNT1262b2 | 0.87 (0.69-1.11) | 0.86 (0.62-1.18) | 0.47 (0.06-3.67) | 1.60 (0.81-3.17) | 1.08 (0.80-1.46) | 0.94 (0.71-1.25) | 1.00 (0.72-1.41) | 1.67 (0.23-12.10) | 1.77 (0.84-3.74) | 1.12 (0.76-1.64) |
| Non-infected | 1 (ref) | 1 (ref) | 1 (ref) | 1 (ref) | 1 (ref) | 1 (ref) | 1 (ref) | 1 (ref) | 1 (ref) | 1 (ref) |
| SARS-CoV-2 infection | 0.97 (0.75-1.26) | 1.01 (0.86-1.20) | 0.98 (0.73-1.31) | 0.83 (0.56-1.22) | 1.20 (0.73-1.99) | 1.03 (0.84-1.27) | 1.05 (0.93-1.19) | 1.02 (0.79-1.30) | 0.98 (0.83-1.16) | 1.16 (1.04-1.30) |
| **MS** |  |  |  |  |  |  |  |  |  |  |
| Unvaccinated | 1 (ref) | 1 (ref) | 1 (ref) | 1 (ref) | 1 (ref) | 1 (ref) | 1 (ref) | 1 (ref) | 1 (ref) | 1 (ref) |
| 1^st^ dose BNT162b2 | 2.05 (1.11-3.78) | 0.77 (0.53-1.12) | NA | 1.73 (0.50-5.96) | 0.95 (0.65-1.39) | 1.58 (0.92-2.70) | 0.72 (0.53-0.99) | NA | 1.60 (0.84-3.06) | 0.94 (0.70-1.26) |
| 2^nd^ dose BNT162b2 | 1.83 (0.50-6.69) | 0.44 (0.21-0.93) | NA | 0.24 (0.05-1.23) | 0.75 (0.38-1.48) | 1.55 (0.47-5.13) | 0.43 (0.13-1.40) | NA | 0.32 (0.06-1.58) | 0.81 (0.33-2.01) |
| 3^rd^ dose BNT1262b2 | 0.26 (0.03-1.97) | 0.30 (0.12-0.72) | NA | NA | 0.23 (0.09-0.54) | 0.67 (0.13-3.41) | 0.35 (0.14-0.93) | NA | NA | 0.40 (0.17-0.97) |
| Non-infected | 1 (ref) | 1 (ref) | 1 (ref) | 1 (ref) | 1 (ref) | 1 (ref) | 1 (ref) | 1 (ref) | 1 (ref) | 1 (ref) |
| SARS-CoV-2 infection | 1.60 (0.52-4.91) | 0.94 (0.49-1.78) | NA | 0.28 (0.02-5.13) | 0.93 (0.47-1.85) | 0.41 (0.03-5.05) | 1.12 (0.58-2.13) | NA | 0.44 (0.02-10.05) | 0.99 (0.55-1.81) |
| **Type 1 diabetes** |  |  |  |  |  |  |  |  |  |  |
| Unvaccinated | 1 (ref) | 1 (ref) | 1 (ref) | 1 (ref) | 1 (ref) | 1 (ref) | 1 (ref) | 1 (ref) | 1 (ref) | 1 (ref) |
| 1^st^ dose BNT162b2 | 1.08 (1.02-1.14) | 1.11 (1.01-1.23) | 0.93 (0.86-1.02) | 1.19 (0.95-1.50) | 1.20 (1.08-1.33) | 1.02 (0.94-1.10) | 1.05 (0.96-1.14) | 0.92 (0.82-1.04) | 1.11 (0.92-1.35) | 1.05 (1.00-1.11) |
| 2^nd^ dose BNT162b2 | 1.06 (1.00-1.12) | 1.05 (0.98-1.12) | 0.88 (0.74-1.04) | 1.22 (1.06-1.41) | 1.17 (1.03-1.34) | 0.99 (0.90-1.10) | 0.99 (0.94-1.05) | 0.92 (0.79-1.09) | 1.09 (0.97-1.23) | 0.96 (0.90-1.03) |
| 3^rd^ dose BNT1262b2 | 0.96 (0.83-1.12) | 0.90 (0.73-1.11) | 0.91 (0.11-7.31) | 1.11 (0.67-1.84) | 1.09 (0.89-1.34) | 0.86 (0.71-1.04) | 0.82 (0.66-1.03) | 0.86 (0.12-6.16) | 1.13 (0.98-1.29) | 0.78 (0.67-0.91) |
| Non-infected | 1 (ref) | 1 (ref) | 1 (ref) | 1 (ref) | 1 (ref) | 1 (ref) | 1 (ref) | 1 (ref) | 1 (ref) | 1 (ref) |
| SARS-CoV-2 infection | 0.98 (0.89-1.07) | 1.03 (0.96-1.10) | 0.95 (0.88-1.02) | 0.97 (0.89-1.07) | 1.16 (0.96-1.39) | 0.94 (0.87-1.01) | 1.00 (0.94-1.05) | 0.92 (0.87-0.98) | 0.97 (0.92-1.03) | 1.04 (0.91-1.20) |
